# Computable Phenotypes for Respiratory Viral Infections in the *All of Us* Research Program

**DOI:** 10.1101/2025.01.17.25320744

**Authors:** Bennett J Waxse, Fausto Andres Bustos Carrillo, Tam C Tran, Huan Mo, Emily E Ricotta, Joshua C Denny

**Affiliations:** National Human Genome Research Institute, National Institutes of Health, Bethesda, MD, USA; National Institute of Allergy and Infectious Diseases, National Institutes of Health, Bethesda, MD, USA; Department of Preventive Medicine and Biostatistics, Uniformed Services University of the Health Sciences, Bethesda, MD, USA; All of Us Research Program, National Institutes of Health, Bethesda, MD, USA

**Author notes:** **Corresponding author:** Josh Denny, MD, MS.

**Keywords:** Electronic Health Records, Phenotype, Respiratory Tract Infections, Reproducibility of Results, Precision Medicine

## Abstract

Electronic health records (EHRs) contain rich temporal data about infectious diseases, but an optimal approach to identify infections remains undefined. Using the *All of Us* Research Program, we developed computable phenotypes for respiratory viruses by integrating billing codes, prescriptions, and laboratory results within 90-day episodes. Phenotypes computed from 265,222 participants yielded cohorts ranging from 238 (adenovirus) to 28,729 (SARS-CoV-2) cases. Virus-specific billing codes showed varied sensitivity (8-67%) and high positive predictive value (90-97%), except for influenza virus and SARS-CoV-2 where lower PPV (69-70%) improved with increasing billing codes. Identified infections exhibited expected seasonal patterns and virus proportions when compared with CDC data. This integrated approach identified episodic disease more effectively than individual components alone and demonstrated utility in identifying severe infections. The method enables large-scale studies of host genetics, health disparities, and clinical outcomes across episodic diseases.

## INTRODUCTION

Respiratory infections are among the most common human diseases. Severity is influenced by demographics, social determinants of health, comorbidities, immunosuppression, lifestyle, exposures, and genetic factors.^1^ Influenza virus, respiratory syncytial virus (RSV), and SARS-CoV-2 are well-studied causes of lower respiratory tract infections, but over 25 known viruses can cause such disease.^2^ The advent of multiplex testing has revealed that rhinovirus (RV), human metapneumovirus (hMPV), parainfluenza viruses (PIV), and common human coronaviruses (hCoVs) can cause similar symptoms, detection rates, morbidity, mortality, and healthcare costs in hospitalized patients.^3–5^

Risk factors for severe infection by influenza virus, RSV, and SARS-CoV-2 are well characterized. These include age extremes, immunosuppression, male sex, smoking, and comorbidities such as obesity, chronic lung disease, hypertension, diabetes, and heart failure.^1,6–8^ Similarly, population-level genetic risk factors have been identified through genome-wide studies for SARS-CoV-2, avian influenza virus (H7N9), pandemic influenza virus (H1N1_pdm09_), and influenza virus by survey report.^9–14^ While underrecognized viruses share similar risk factors like advanced age and immunocompromised status, they remain understudied.^3,4,15,16^

Population-level studies of respiratory infections typically rely on administrative claims data, laboratory surveillance data, or curated clinical cohorts.^15,17^ While laboratory results and some pathogen-specific billing codes are highly specific, they have poor sensitivity due to infrequent testing, and adding non-specific International Classification of Diseases (ICD) codes only modestly improves sensitivity.^18,19^ In contrast, electronic health record (EHR)-based phenotyping algorithms can integrate multiple data types to reliably identify disease cohorts for observational studies. Although integrating billing codes, clinical notes, and medications in disease phenotyping using EHRs improves performance, this approach has only rarely been used for respiratory viruses.^15,20–23^

This study aimed to create and assess a computable phenotype for identifying viral respiratory infections in EHR data using the National Institutes of Health’s *All of Us* Research Program (*All of Us*). We evaluated ICD billing codes, medications, and laboratory results to identify infections, and assessed phenotype performance through specificity, positive predictive value (PPV) and sensitivity calculations compared to gold-standard laboratory testing; and compared the frequency and distribution of laboratory results against CDC surveillance data.

## MATERIALS AND METHODS

### Data Acquisition

We analyzed data from the *All of Us* Research Program, which digitally enrolls participants aged 18 years and older across the United States.^24^ *All of Us* is a large, diverse national cohort where participants contribute survey data, standardized physical measurements, biospecimens, and EHR data including billing codes, prescriptions, and laboratory results [23]. Participants provide consent to share health information, which includes physical measurements, surveys, genomic data, and EHRs. The informed consent and enrollment process has been described, and specific Institutional Review Board approval is not required for Controlled Tier use of de-identified data, deemed nonhuman subjects research by the *All of Us* Institutional Review Board.^24,25^ The program prioritizes recruitment of populations historically underrepresented in biomedical research.^24^ This analysis used Controlled Tier data (C2022Q4R13) from the *All of Us* Researcher Workbench and was restricted to the 265,222 participants who had ICD codes, medications, or laboratory results in their EHR data between 1/1/1981 and 7/1/2022. Data linkage, follow-up completeness, quality assessment, privacy, and community engagement are described in the *All of Us* Protocol.^26^ This study meets all five of the CODE-EHR minimum framework standards for the use of structured health care data in clinical research, with one out of five standards meeting preferred criteria.^27^ Participants’ demographic data were derived from the *All of Us* Researcher Workspace’s “person” table and its “The Basics” survey.

### Phenotype Development

We developed computable phenotypes for eight respiratory viruses: rhinovirus (RV); human metapneumovirus (hMPV); respiratory syncytial virus (RSV); adenovirus (ADV); SARS-CoV-2, parainfluenza (PIV); common human coronavirus (hCoV); and influenza virus. Patient encounters were identified in the EHR if they had at least one of the following: a virus-specific billing code (ICD-9-CM or ICD-10-CM), an antiviral indicated for the target pathogen, or a positive laboratory test. We identified virus-specific billing codes and Logical Observation Identifiers Names and Codes (LOINC) laboratory results by searching for the virus name and related terms (*e.g.*, “adenovirus” and “adenoviral pneumonia”). We excluded codes and results for zoonotic infections, vaccine-related events, and ICD codes for explicitly non-respiratory conditions (*e.g.*, “ovine adenovirus”, “enteritis due to adenovirus”). We also excluded codes and results for pathogens sharing components of the virus name (*e.g.*, “Haemophilus influenzae”). Laboratory data included nucleic acid amplification, antigen, and culture results. For influenza virus and SARS-CoV-2, we included antiviral medications (*i.e.*, oseltamivir, zanamivir, and baloxavir for influenza virus; remdesivir, molnupiravir, and nirmatrelvir/ritonavir for SARS-CoV-2) given for more than one day. For oseltamivir and zanamivir, prescriptions were removed if the duration of treatment was greater than 6 days to exclude prophylaxis. Antivirals for other respiratory viruses were not included due to poor specificity or their reserved use for severe or immunocompromised cases (Figure 1A).

**Figure 1.**
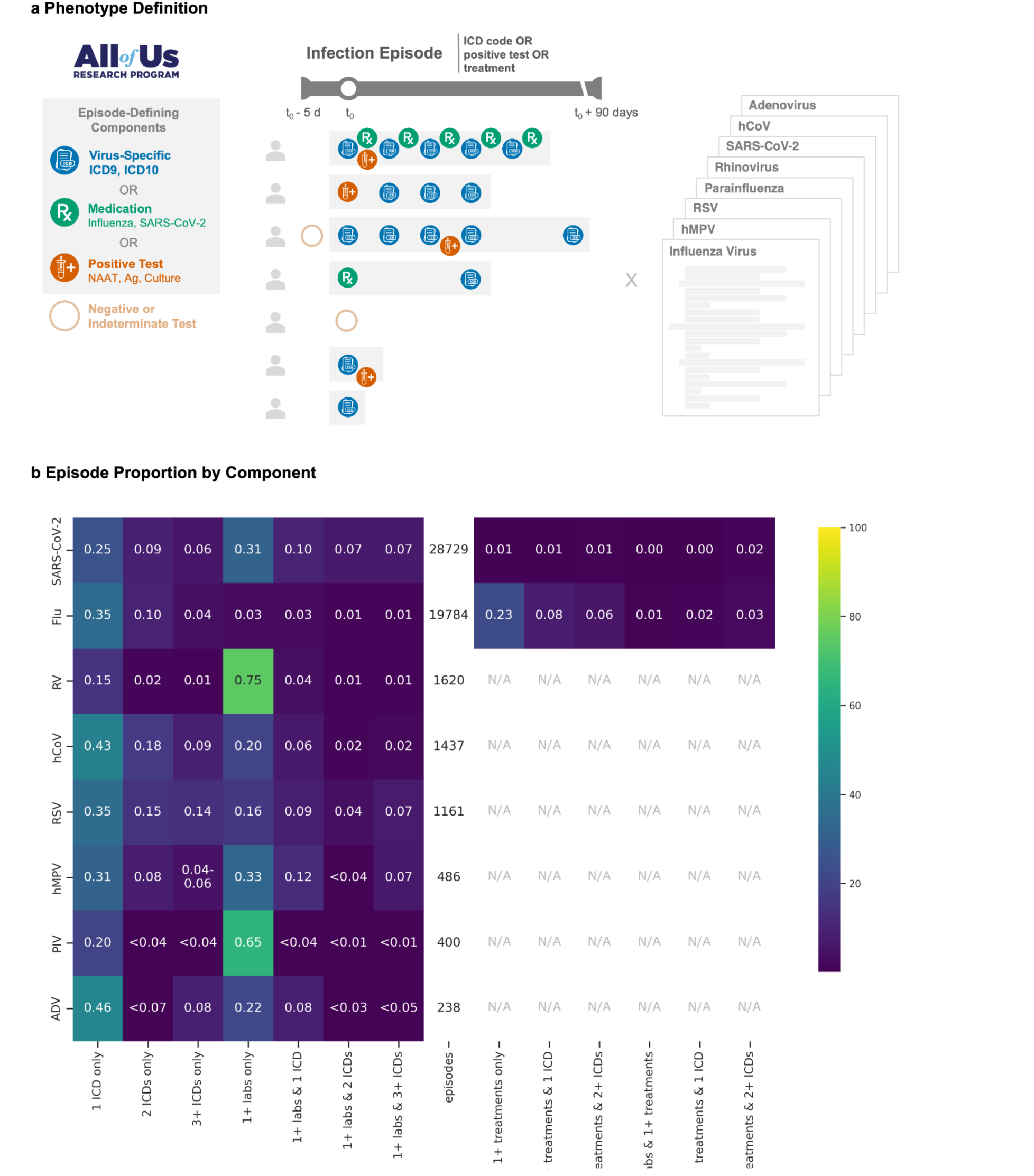
Computational phenotype for respiratory virus episodes using electronic health records (EHRs) with respiratory episode composition. A: Episodes were defined by (1) virus-specific ICD-9-CM or ICD-10-CM codes, (2) antiviral medications (for influenza and SARS-CoV-2), and/or (3) positive laboratory results including nucleic acid amplification tests (NAAT), antigen tests, or cultures. The first qualifying event is designated as time zero (t_0_), and all related, subsequent events within 90 days were grouped into the same episode. Negative or indeterminate tests were also included, with a five-day lookback window to incorporate false negative results. Phenotypes were computed for influenza, human metapneumovirus (hMPV), respiratory syncytial virus (RSV), parainfluenza, rhinovirus (RV), SARS-CoV-2, common human coronavirus (hCoV), and adenovirus (ADV). Heatmap (B) showing the breakdown of episode types (columns) for each virus (row). Colors indicate the percentage of counts for each virus (*e.g.*, 74.9% of RV episodes (1,210/1,620) contained only positive laboratory results. Percentage and corresponding color for counts below 20 (with a second count to prevent back-calculation for hMPV), were censored per the *All of Us* participant privacy policy. N/A : not applicable.

We computed phenotypes for each infection episode by grouping related clinical events (Figure 1A). An episode began with the first occurrence (t_0_) of any virus-specific component: a virus-specific ICD code, positive laboratory result, or qualifying antiviral prescription. All subsequent components within 90 days of t_0_ were considered part of the same episode, while events beyond 90 days initiated new episodes.^15^ To capture false negatives, we included negative or indeterminate laboratory results from t_0_-5 days through the episode’s end. The per-episode occurrence of constituent components (ICD codes, positive laboratory results, and medications) were tallied (Figure 1B). Counts for all virus-specific ICD codes, laboratory results, and medications used for phenotyping are provided (Tables S1-3). We de-duplicated row-level entries; categorized visit types into major categories (*i.e.*, intensive care unit, inpatient, emergency room, urgent care, post-acute care, outpatient, and unknown); and reclassified laboratory results as positive, negative, or indeterminate (Table S4).

### Phenotype Sensitivity Analyses

#### Positive Predictive Value, Specificity, and Sensitivity Calculations

Using non-antigen test results as reference standard, we calculated the performance (PPV, sensitivity, and specificity) based on increasing counts of virus-specific ICD codes within each episode (*e.g.*, multiple occurrences of J10.1, or combinations like J10.1 + J11 + 487). For a given threshold *N* (n = 0, n ≥ 1, n ≥ 2, n ≥ 3, n ≥ 4), we defined true positives as episodes with *N* virus-specific ICD codes and a positive test, false positives as episodes with *N* ICD codes and only negative tests, and false negatives as episodes with fewer than *N* ICD codes and a positive test.

For influenza virus and SARS-CoV-2, we additionally tested the performance of incorporating antiviral prescriptions. We first assessed specificity, sensitivity, and PPV in cases requiring both the specified ICD count and a prescription to be considered positive, or fewer than *N* ICD codes and no prescription to be considered negative. Then, we evaluated full performance metrics (sensitivity, specificity, PPV, negative predictive value (NPV), and phi coefficient) across different combinations of ICD code thresholds and medication criteria for these two viruses.

#### Temporal and Geographic Analysis of Phenotype-Positive Episodes

For any episode that met positivity criteria, we analyzed temporal patterns by calculating three-week moving averages of episode and constituent component counts from July 2017 (MMWR week 201726) through June 2022 (MMWR week 202225). Because hCoV PPV was lower than expected for non-influenza, non-SARS-CoV-2 viruses, and as hCoV ICD counts were disrupted during the COVID-19 pandemic, hCoV ICD codes after February 1, 2020 were excluded from the analysis (Figure S3). We assessed the geographic distribution of episode rates by three-digit ZIP code prefixes (zip3).

#### Level of Care Sensitivity Analysis

Because of differences in testing and care by facility type and acuity, for each episode we identified the highest acuity encounter (from lowest to highest: outpatient, post-acute care, urgent care, emergency department, or inpatient) within a window spanning t_0_-7 days through t_0_+14 days. We chose this window after analyzing the distributions of visit timing for phenotype-related visits (those associated with virus-specific ICD codes, antivirals, or laboratory results) and all visits (Figure S4). Use of all visits within the selected window, rather than only phenotype-related visits, reduced the overall percentage of missing encounter data from 25.9% to 15.4%; across viruses and ICD counts, median missing encounter data decreased from 17.6% (IQR 7.6-28.3%) to 11.3% (IQR 2.2-17.8%) (Table S5). Rarely (0.09%-0.43%), phenotype-related maximum encounter acuity exceeded encounters within this time window (*e.g.*, hospitalization occurring >7 days prior to initial viral diagnosis, Figure S5). For PIV, hMPV, and RSV, manual review of these visits revealed that the associated encounter start date preceded the window period by a few days, and the phenotype component level of care was retained. We analyzed level-of-care patterns across viruses, stratifying by ICD codes and test positivity.

### Comparison with National Surveillance Data

To understand the representativeness of respiratory illness data in *All of Us,* we compared the seasonal percent positivity and test volume of *All of Us* EHR laboratory results to data from three CDC/WHO sources: National Respiratory and Enteric Virus Surveillance System (NREVSS), COVID-19 Data Tracker, and the Global Influenza Surveillance and Response System (GISRS).

First, we assessed geographic coverage by comparing *All of Us* participant locations and testing rates to deduplicated NREVSS clinical laboratory results from 2017-2021.^28^ *All of Us* EHR participant counts were visualized by aggregating data across three-digit ZIP code prefixes (zip3). Zip3 information was missing for 2/265,222 (0.00%). *All of Us* EHR participants (per 1,000 zip3 2020 Census population) and *All of Us* participants tested (per 1,000 *All of Us* participants with EHR data) were similarly aggregated by zip3 code. Zip3 regions with five or fewer *All of Us* participants were removed and classified as “No Data.” We obtained zip3 boundaries from US Census Bureau zip code tabulation areas, 2017 cartographic state boundaries from the Vega us-10m.json dataset, and 2020 zip code tabulation area populations from the US Census Bureau.

Next, we assessed virus distribution by comparing proportions detected (% = N type / N total with known type) in *All of Us* to surveillance data from NREVSS and the GISRS.^29–31^ These comparisons were limited to hCoV, influenza virus, and PIV, as these were the only viruses for which syndromic multiplex panels routinely report type-specific results.

To evaluate temporal patterns, we compared weekly test positivity data between *All of Us* and CDC surveillance from the first week of July, 2016 to the last week of June, 2022. For each virus, we calculated the percentage of positive tests for each *MMWR* reporting week. We plotted three-week moving averages for both percent positivity and total tests performed for *All of Us* and CDC data. We obtained CDC comparison data from NREVSS for non-SARS-CoV-2 viruses, additional influenza virus data from FluView, and SARS-CoV-2 data from the COVID Data Tracker.^28,32,33^

## RESULTS

### Cohort Characteristics

Among 265,222 *All of Us* participants with ICD codes, medication entries, or laboratory results recorded between 1/1/1981 and 7/1/2022, we identified respiratory virus episodes that varied substantially in duration and composition (Figure 1B). SARS-CoV-2 (n [distinct episodes]=28,729) and influenza virus (n=19,784) were the largest cohorts, followed by RV (n=1,620), hCoV (n=1,437), and RSV (n=1,161) and the smallest cohorts, hMPV (n=486), PIV (n=400), and ADV (n=238).

Across all cohorts, participants were predominantly female (61-68%) with median ages mostly between 50 and 58 (Table S6). Participants who self-reported as White were the plurality for every virus (32.9-60.1%), compared to participants self-reporting as Black (16.5-28.5%) or Hispanic/Latino (17-32.1%). All other options (Asian, multiple selected, Middle Eastern or North African, and Native Hawaiian or Other Pacific Islander) were rare (0-2.2%). SARS-CoV-2 and influenza virus participant demographics most closely mirrored the overall *All of Us* cohort with ICD, laboratory, or medication data. These two groups more frequently self-reported as White (50.4-60.1%) and were more frequently employed with higher reported income, education, and employer-provided insurance. Demographic data were only notably missing for insurance type (46,487/265,222=17.5% for all participants with EHR data). Test count per person was higher among infected cohorts than tested cohorts for each virus.

Episodes commonly consisted of either laboratory results alone (predominant for RV [74.7%], PIV [65.0%], SARS-CoV-2 [31.3%], hMPV [32.9%]) or single ICD codes (predominant for ADV [45.8%], hCoV [43.1%], RSV [35.4%], influenza [34.8%]) (Figure 1B). Antiviral use varied markedly: SARS-CoV-2 episodes rarely included antiviral prescriptions (4.57%), while medication-only episodes were frequently observed for influenza virus (22.9%), even after excluding prophylactic prescriptions.

### Phenotype Performance for Detecting True Positives

To understand the diagnostic performance of varying ICD codes for an episode, we calculated sensitivity, specificity, and PPV for each virus across *N* ICD codes per episode using non-antigen test results as a reference standard (Figure 2). The sensitivity of using one or more ICD codes varied between viruses and decreased as the minimum *N* ICD codes increased. Diagnoses using one or more ICD codes for influenza virus had the highest sensitivity (66.8%), compared to moderate sensitivity for RSV (55.2%), SARS-CoV-2 (44.8%), ADV (42.4%), hMPV (40.2%), and hCoV (33.4%), and minimal sensitivity for RV (9.2%) and PIV (8.3%).

**Figure 2.**
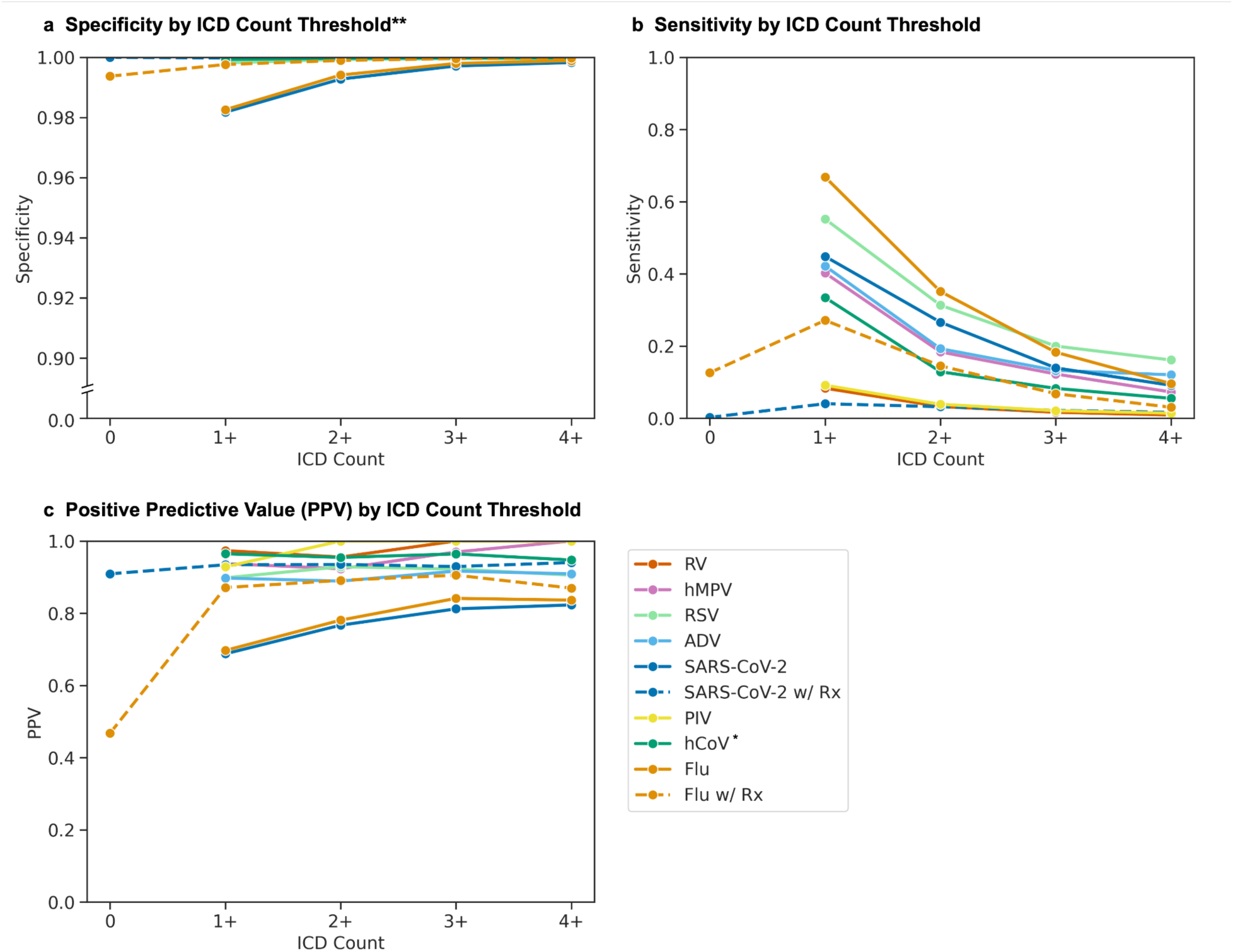
Phenotype performance across different episode definitions. Specificity (A), sensitivity (B), and PPV (C) were calculated using non-antigen laboratory results as the reference standard. For each virus, colored lines show performance across episodes containing increasing numbers of ICD codes. For influenza virus (dashed orange line) and SARS-CoV-2 (dashed dark blue line), additional lines show performance when episodes were restricted to episodes containing both ICD codes and antiviral prescriptions (Rx). *Human coronavirus (hCoV) episodes excluded ICD codes after February 1, 2020, due to loss of specificity during the COVID-19 pandemic (Figure S3A). **Y-axis for specificity (A) is broken to depict differences near 1.0. PIV: parainfluenza; hMPV: human metapneumovirus; RV: rhinovirus; ADV: adenovirus; RSV: respiratory syncytial virus; hCoV: human coronavirus; Flu: influenza virus.

Specificity and PPV demonstrated similar patterns, with exaggerated variation in PPV initially demonstrating three groupings. First, for influenza virus and SARS-CoV-2, the PPV for one or more ICD codes was lower (69.7% and 68.8%, respectively), but increased as minimum *N* ICD count increased (78.1% and 76.7% for at least 2 ICD codes, respectively) (Figure 2). Second, for non-influenza, non-SARS-CoV-2 viruses, the PPV was high and did not change substantially as ICD count increased (89.7-97.3%). Third, hCoV initially demonstrated a high PPV (79.5%) that decreased as ICD count increased (71.8% for at least 2 ICD codes) (Figure S3A).

During the COVID-19 pandemic, hCoV ICD code counts spiked above historical maxima despite an absence of positive tests (Figure S3B). After removing hCoV ICD codes after February 1, 2020, the PPV trend for common hCoV became similar to other non-influenza, non-SARS-CoV-2 viruses (Figure 2, Figure S3A).

Adding medication use to the phenotype had varying effects on performance. As with the medication-exclusive phenotypes, specificity and PPV increased with each additional ICD code for the medication-inclusive influenza and SARS-CoV-2 cohorts. While only a small proportion (1,345/28,741=4.67%) of SARS-CoV-2 episodes included a prescription for remdesivir, molnupiravir or nirmatrelvir, the addition of medication to the phenotype did increase PPV for this subset of 1,345 participants (Figure 2). For influenza virus, medication use alone was poorly predictive (PPV=46.8%), but combining medications with 1 ICD code improved PPV compared to 1 or more codes alone (87.1% vs. 69.7%, respectively) (Figure 2).

Varying ICD code thresholds and antiviral requirements in the entire influenza and SARS-CoV-2 cohorts demonstrated an expected trade-off in performance (Table 1). For both viruses, at least one ICD code or medication was the most sensitive phenotype (76.0% influenza virus and 45.1% SARS-CoV-2), but this caused the highest number of false positives and the lowest PPVs (65.8% and 68.8%, respectively). For influenza virus, by requiring at least two ICD codes or a medication accompanied by an ICD code, the lower sensitivity (47.7%) was accompanied by a marked reduction in false positives (778 to 238) and increase in PPV (65.8% to 79.8%). Similar trends were observed for SARS-CoV-2, and despite trade-offs, the phi coefficient was highest for the broadest phenotypes.

**Table 1.** Phenotype Performance

| | Phenotype | TP | Counts (N) | | TN | Sen. | Phenotype Performance | | | $\phi$ |
| --- | --- | --- | --- | --- | --- | --- | --- | --- | --- | --- |
|  |  |  | FP | FN |  |  | Spec. | PPV | NPV |  |
| Flu | 1+ ICDs or medication | 1496 | 778 | 472 | 32018 | 0.760 | 0.976 | 0.658 | 0.985 | 0.688 |
|  | 1+ ICDs | 1315 | 572 | 653 | 32224 | 0.668 | 0.983 | 0.697 | 0.980 | 0.664 |
|  | 2+ ICDs or medication | 1120 | 444 | 848 | 32352 | 0.569 | 0.986 | 0.716 | 0.974 | 0.619 |
|  | 2+ ICDs or (1 ICD + medication) | 939 | 238 | 1029 | 32558 | 0.477 | 0.993 | 0.798 | 0.969 | 0.600 |
|  | 3+ ICDs or medication | 941 | 339 | 1027 | 32457 | 0.478 | 0.990 | 0.735 | 0.969 | 0.574 |
|  | 3+ ICDs or (1-2 ICDs + medication) | 760 | 133 | 1208 | 32663 | 0.386 | 0.996 | 0.851 | 0.964 | 0.558 |
|  | 2+ ICDs | 691 | 194 | 1277 | 32602 | 0.351 | 0.994 | 0.781 | 0.962 | 0.506 |
|  | 3+ ICDs | 360 | 68 | 1608 | 32728 | 0.183 | 0.998 | 0.841 | 0.953 | 0.379 |
| COVID | 1+ ICDs or medication | 7298 | 3302 | 8894 | 177219 | 0.451 | 0.982 | 0.688 | 0.952 | 0.526 |
|  | 1+ ICDs | 7258 | 3298 | 8934 | 177223 | 0.448 | 0.982 | 0.688 | 0.952 | 0.524 |
|  | 2+ ICDs or medication | 4483 | 1324 | 11709 | 179197 | 0.277 | 0.993 | 0.772 | 0.939 | 0.438 |
|  | 2+ ICDs or (1 ICD + medication) | 4443 | 1320 | 11749 | 179201 | 0.274 | 0.993 | 0.771 | 0.938 | 0.435 |
|  | 3+ ICDs or medication | 2598 | 546 | 13594 | 179975 | 0.160 | 0.997 | 0.826 | 0.930 | 0.345 |
|  | 3+ ICDs or (1-2 ICDs + medication) | 2558 | 542 | 13634 | 179979 | 0.158 | 0.997 | 0.825 | 0.930 | 0.342 |
|  | 2+ ICDs | 4309 | 1310 | 11883 | 179211 | 0.266 | 0.993 | 0.767 | 0.938 | 0.427 |
|  | 3+ ICDs | 2260 | 523 | 13932 | 179998 | 0.140 | 0.997 | 0.812 | 0.928 | 0.318 |

TP: true positive; FP: false positive; FN: false negative; TN: true negative; Sen: sensitivity; Spec: specificity; PPV: positive predictive value; NPV: negative predictive value; $\phi$ : phi coefficient (mean square contingency coefficient).

We identified infection episodes nationwide, with SARS-CoV-2 and influenza virus demonstrating the broadest US coverage. While episodes generally matched the *All of Us* EHR subgroup distribution (Figure S1C), incorporating ICD codes and medications exhibited higher infection rates in the Southeast and Texas despite lower testing coverage in these regions (Figure 3). Temporally, episodes composed of 1-3 ICD codes showed seasonality patterns consistent with test-positive episodes for all frequently detected viruses (Figure 4). During the early COVID-19 pandemic (winter 2020 to spring 2021), only SARS-CoV-2 and RV were consistently identified.

**Figure 3.**
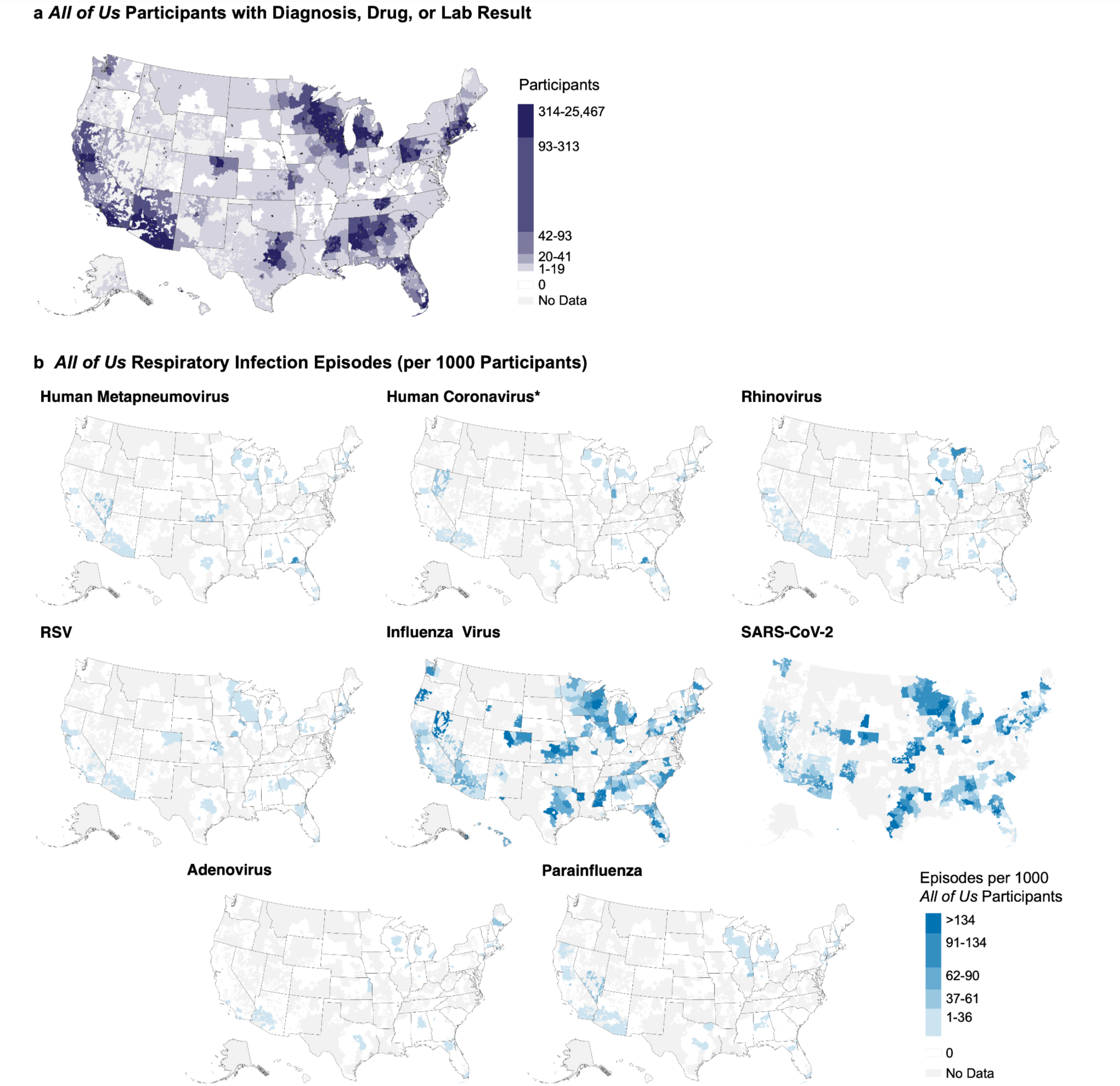
Geographic distribution of respiratory virus episodes. Heat maps show episode rates per 1,000 *All of Us* participants with EHR data by three-digit zip code prefix. Colors represent quintiles defined by SARS-CoV-2 rates, the largest cohort. Regions with five or fewer *All of Us* participants are marked as “No Data” in (B). *Human coronavirus episodes were filtered as described in methods. RSV: respiratory syncytial virus.

**Figure 4.**
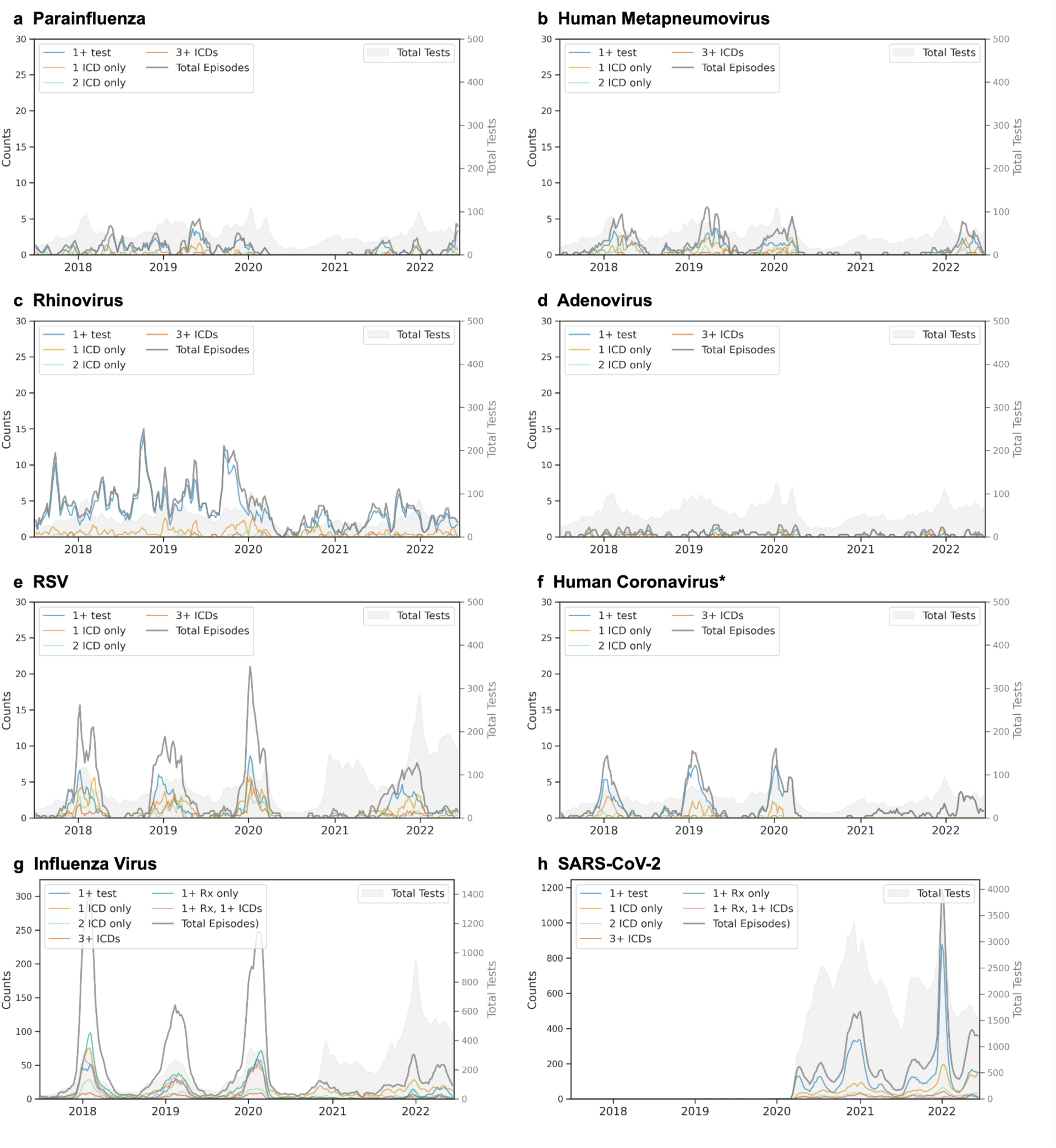
Temporal patterns of respiratory virus episodes by composition. Three-week moving averages shown for (A) parainfluenza, (B) human metapneumovirus, (C) rhinovirus, (D) adenovirus, (E) respiratory syncytial virus (RSV), (F) human coronavirus, (G) influenza virus, and (H) SARS-CoV-2. For each virus, lines show total episodes (gray) and episodes by composition: one or more positive tests (blue), single ICD code (light orange), two ICD codes (light green), and three or more ICD codes (red-orange). For influenza, and SARS-CoV-2, additional lines show episodes with antiviral prescriptions alone (dark green) or with concurrent ICD codes (pink). Gray shading indicates testing volume. Left y-axis corresponds to episode counts; right y-axis shows total tests performed. *Episodes for hCoV are shown after applying temporal filtering (unfiltered plot compared in Figure S3).

### Patterns in Phenotype Composition by Level of Care

Encounter level of care varied by virus and episode composition. For RV, hMPV, PIV, hCoV, and SARS-CoV-2, episodes defined by at least one test without ICD codes were the most frequent, while for RSV, ADV, and influenza virus, ICD-only episodes predominated (Figure 5). Influenza virus episodes with antiviral prescriptions showed a similar distribution of visit types compared to those without, while SARS-CoV-2 episodes rarely included prescriptions during our study period.

**Figure 5.**
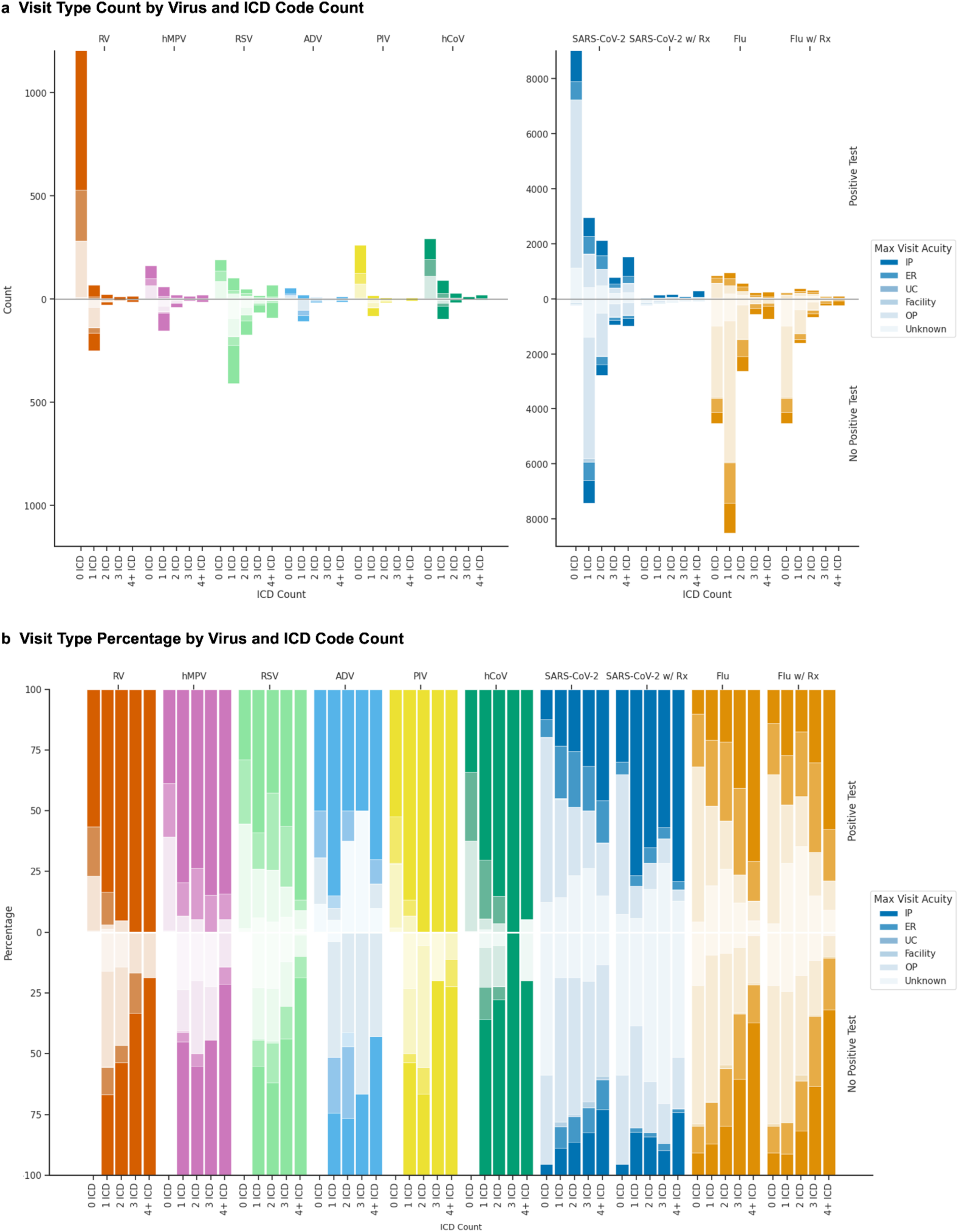
Level of care patterns by virus and episode composition. For each virus, (A) counts and (B) percentages of maximum level of care recorded between seven days before and 14 days after episode start. Results are stratified by episode characteristics: episodes with positive tests (upper panels) versus those without (lower panels), and by number of ICD codes (columns). Color intensity corresponds to level of care: inpatient (IP, darkest), emergency room (ER), urgent care (UC), outpatient (OP), and unknown (lightest). For influenza virus (Flu) and SARS-CoV-2, additional columns show episodes containing antiviral prescriptions. PIV: parainfluenza; hMPV: human metapneumovirus; RV: rhinovirus; ADV: adenovirus; RSV: respiratory syncytial virus; hCoV: human coronavirus; Rx: antiviral prescription.

By percentage, influenza virus and SARS-CoV-2 episodes showed a mix of outpatient, ER, and inpatient encounters, while other viruses demonstrated higher rates of ER visits and hospitalization. Episodes containing a positive test consistently showed higher rates of hospitalization compared to test-negative episodes. Similarly, hospitalization rates increased as the number of ICD codes within an episode increased. The cohorts included very few post-acute care encounters and almost no urgent care encounters.

### Laboratory Result Comparison

Using national epidemiological data from NREVSS, COVID Data Tracker, and GISRS, we compared *All of Us* laboratory results by geographic coverage, virus type proportion, and temporal trends.

We found broad US coverage of *All of Us* participants with relevant EHR data (265,222 participants), with enriched sampling near population centers in the Northeast megalopolis; Western Pennsylvania; Great Lakes Region; Southeast; Arizona; California; and the metropolitan areas of Austin/Dallas, Kansas City, Denver, and Seattle (Figure S1A, Table S7). Only 3.7% of zip3 codes had no *All of Us* participants with ICD, laboratory, or medication data. Testing patterns in the *All of Us* data overlapped with CDC clinical laboratories reporting to NREVSS (Figure S1B) and mirrored participant distribution (Figure S1A) with a notable decrease in testing for all respiratory viruses in the Southeast relative to participant density (Figure S1C). Testing frequency varied substantially by virus; participants were more frequently tested for influenza virus and SARS-CoV-2 compared to all other viruses.

Virus type distributions in *All of Us* were similar to national surveillance data from NREVSS and GISRS.^29–31^ For PIV (2011-2019), HPIV-3 was most commonly detected and all other types were less frequent (Figure S2A). For hCoV (2014-2021), OC43 was most common and 229E was least common, while the order of NL63 and HKU1 differed (Figure S2B). Influenza virus type proportions (2010-2020) were nearly identical, with influenza virus A more common than influenza virus B (Figure S2C). Cross-dataset influenza subtype comparisons were not available, but in *All of Us*, H3N2 and H1N1 pdm09 were markedly more common than H1N1 and H5N1, as expected.

Test positivity patterns from 2017 to 2022 matched CDC rates for most viruses (mean absolute error 5.89 percent positive tests per week for RV and 1.18-2.82 for all other viruses) (Figure 6). SARS-CoV-2, influenza virus, and RV showed the highest percent positivity, and most viruses showed expected seasonal patterns: PIV and RV exhibited two seasonal peaks per year (spring-dominant for PIV, fall-dominant for RV), while RSV, influenza, and hMPV demonstrated single overlapping winter peaks. SARS-CoV-2 positivity matched expected variant waves (*e.g.*, Alpha, Delta, and Omicron BA.1). Notable differences in the *All of Us* data include more variability in RSV tests and positivity, undercounted positivity by ∼10% during peak respiratory season for influenza and RSV, and less ADV positivity, relative to CDC data.

**Figure 6.**
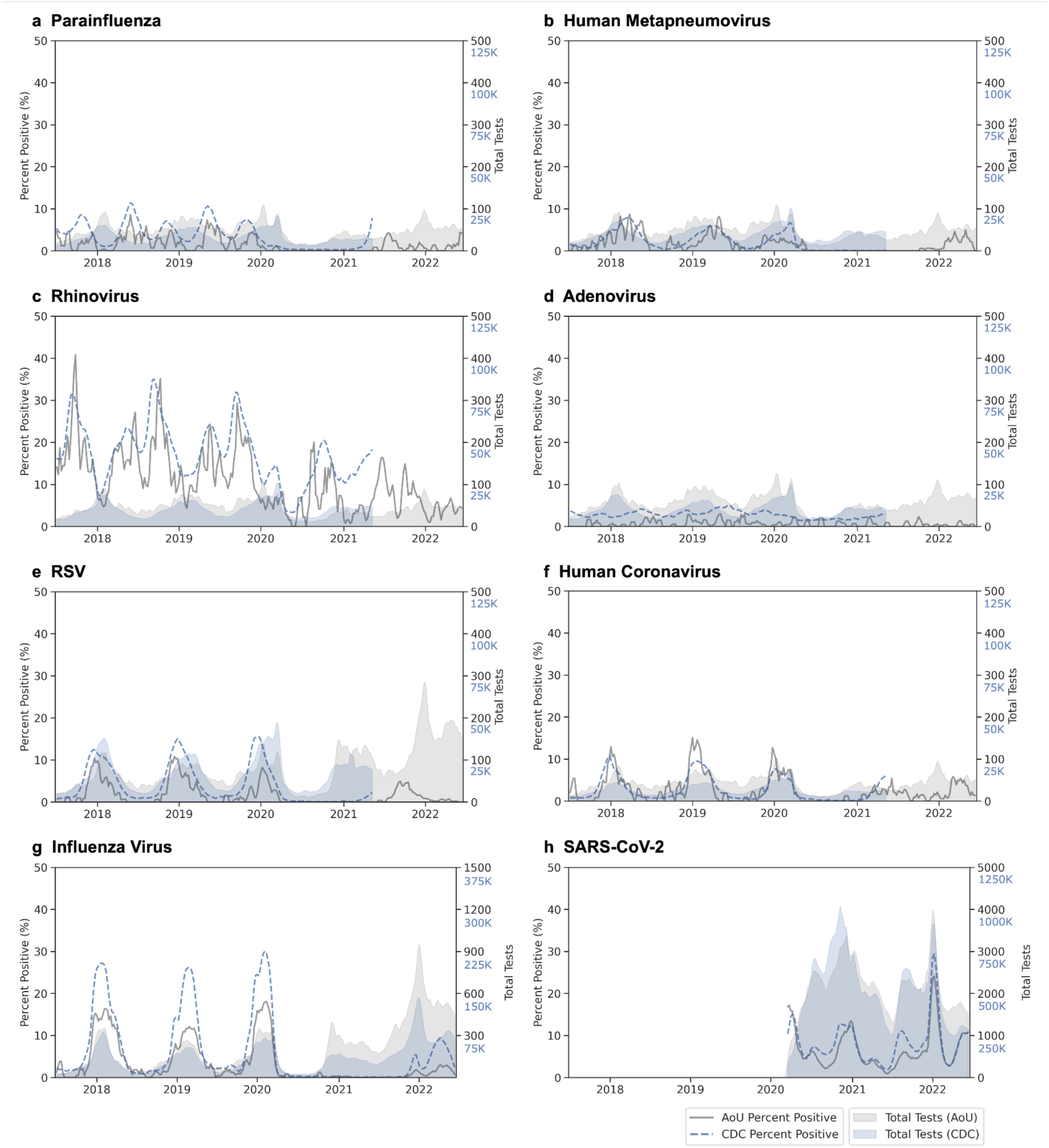
Temporal validation of test positivity using CDC surveillance data. Three-week moving average of test positivity and test volume comparing *All of Us* (gray) to CDC surveillance data (blue) for eight respiratory viruses: (A) parainfluenza, (B) human metapneumovirus, (C) rhinovirus, (D) adenovirus, (E) respiratory syncytial virus (RSV), (F) common human coronavirus (hCoV), (G) influenza virus, and (H) SARS-CoV-2. In each case, percent positivity (lines) corresponds to the left y-axis and total tests performed (shaded areas) corresponds to the right y-axis. CDC data were obtained from NREVSS for panels A-F (ending 2021), FluView for influenza virus, and COVID Data Tracker for SARS-CoV-2. AoU: *All of Us*.

## DISCUSSION

This study demonstrates that combining virus-specific EHR data elements reliably identifies respiratory viral infections in large biobank datasets. Using temporal, geographic, and sensitivity analyses, we described important performance insights into respiratory virus phenotyping. All phenotypes exhibited high specificity, though trade-offs exist between accuracy and sensitivity for influenza virus and SARS-CoV-2.

We identified epidemiological patterns and cohort sizes that matched national surveillance data. The combination of laboratory results, ICD codes, and medications increased case detection beyond what any component alone could identify, which was valuable where laboratory testing was less frequent (e.g., the Southeast). The cohorts varied in size: approximately 20,000-30,000 episodes for influenza virus and SARS-CoV-2 compared to 200-1,600 for other viruses, likely reflecting increased clinical suspicion and testing for common pathogens rather than true differences in disease burden.^15^ These cohorts could be used to investigate host genetic factors, health disparities, geographic and environmental risk factors, and clinical outcomes across respiratory viral infections. The longitudinal available also supports study of lifestyle factors through wearables and patient-reported outcomes from surveys.

Phenotype performance varied substantially between viruses, across episode composition, and over time. Virus-specific ICD code sensitivity varied widely: higher for influenza (66.8% vs. 38-95% in published studies) and RSV (55.2% vs. 24%) and moderate for SARS-CoV-2, ADV, hMPV, and hCoV (33-44%), but very low for PIV (8.3% vs. 14% and RV (9.2% vs. 0%).^18,19,34–36^ While non-influenza, non-SARS-CoV-2 virus-specific ICD codes showed high PPV regardless of count (89.7-97.3%), single ICD codes for SARS-CoV-2 and influenza were less predictive (68.8-69.7%).^18,19,35,36^ For medication-inclusive cases, ICD codes had a higher PPV for influenza virus and SARS-CoV-2 episodes, and for SARS-CoV-2, antivirals alone were highly predictive of test results, although receipt of remdesivir, molnupiravir or nirmatrelvir was rare (4.57% of all episodes) during the study period.

The COVID-19 pandemic disrupted the seasonal transmission of most respiratory viruses, granting the opportunity to assess ICD code performance in unexpected settings and demonstrating the importance of evaluating code performance over time for seasonally variable diseases. hCoV ICD codes were inappropriately used to identify concern for COVID-19 infection, with diminished but persistent effects throughout the study period. Apart from rhinovirus, our phenotypes only rarely identified false positive episodes during the COVID-19 pandemic, mostly attributable to influenza virus episodes composed of a single ICD code. We suspect that these episodes reflect clinical concern for infection rather than true infection, and indeed, adjusting the influenza phenotype from any ICD code or medication to 2+ ICD codes or medications accompanied by an ICD code markedly reduced false positives and increased PPV. In this work, we suggest choosing phenotype characteristics that match the research question and desire to maximize sensitivity vs. PPV.

Level-of-care analyses revealed patterns suggesting systematic detection biases toward higher acuity settings. Our phenotype identified a high frequency of infections at emergency and inpatient visits for RV, hMPV, RSV, ADV, PIV, and hCoV, compared to more outpatient visit types for SARS-CoV-2 and influenza virus. These findings differ from established hospitalization rates: CDC estimates suggest that only 1-2% of medically-attended influenza cases require hospitalization, while COVID-19 hospitalization rates ranged between 2.1% and 68% over this study period, with temporal trends showing a decrease from ∼50% in the early pandemic to 20% by July, 2022.^37–41^ Moreover, prospective studies in adults have shown that other respiratory viruses show either similar or lower hospitalization rates compared to influenza - the opposite of our results.^4,16^ This discordance suggests that the *All of Us* computable phenotype oversamples high levels of care, particularly for non-influenza, non-SARS-CoV-2 viruses, likely due at least three factors: the lack of cost-effective outpatient assays, an absence of specific therapeutic interventions that would justify multiplex testing costs in lower-acuity care settings, and the utility of identifying an etiology in the inpatient setting, where cessation of antibiotics or discharge are considerations.

Several limitations affect the interpretation and generalizability of these findings. The low sensitivity for all viruses indicates that this method cannot be used to study disease prevalence, as nonspecific syndromic coding likely predominates for upper respiratory illnesses. Additionally, the requirement for data conversion to a common data model before release in curated data repositories means this method cannot support real-time surveillance. In addition, while *All of Us* provides broad national coverage, it highly sampled some states (AZ, MA, WI, AL, PA, IL, MI, NY, MS, CA) and left approximately half the US population sampled below rates of 1 in 10,000. The program’s intentional oversampling of populations historically underrepresented in biomedical research, while invaluable to health equity research, also results in demographics that differ from the overall US population. Other known impediments to generalizability compared to the US population include decreased representation of blind and deaf participants; difficulty in linking EHRs from a considerable portion of *All of Us* participants; and the decreased representation of persons of Asian, Middle Eastern or North African, and Native Hawaiian or Other Pacific Islander heritage in this cohort.^42^

Finally, this work faces challenges common to EHR-based research. These include labeling bias, implicit clinician biases which could be influenced by demographics, and informed presence bias where EHR inclusion typically reflects illness rather than routine care.^43,44^ The minimal representation of urgent care and post-acute care in our data further underscore the disconnected nature of healthcare in the US, and identifies a likely gap in infection detection in this cohort.

Despite these limitations, our computable phenotypes reliably detected geographic and temporal patterns of infection matching national surveillance, although severe infections are oversampled and many mild infections are likely missed. This work supports a need for expanded surveillance of non-influenza/non-SARS-CoV-2 pathogens in routine medical care.^45^ Using a cohort that is expected to continually grow, our results enable future studies of genetic susceptibility and clinical outcomes research across both well-studied and understudied respiratory viruses. This work serves as a foundation for the creation and validation of other computable phenotypes for episodic infectious diseases using EHR-based methods.

## DATA AND CODE AVAILABILITY

The Community Workspace “Respiratory Virus Computable Phenotype” is available for all approved *All of Us* users and includes all code and data used in this work (https://workbench.researchallofus.org/workspaces/aou-rw-ae307fda/respiratoryviralinfectionsinallofus/analysis).

## Data Availability

The Community Workspace “Respiratory Virus Computable Phenotype” is available for all approved *All of Us* users and includes all code and data used in this work.

https://workbench.researchallofus.org/workspaces/aou-rw-ae307fda/respiratoryviralinfectionsinallofus/analysis

## ACKNOWLEDGEMENTS

S Olsen and A Winn (CDC) kindly provided percent positivity data and locations of CDC clinical labs reporting to NREVSS. This work received an exception to the Data and Statistics Dissemination Policy from the *All of Us* Resource Access Board for figures 4 and 5. The primary author acknowledges the use of Claude (Anthropic) in the generation and revision of code to produce this computable phenotype, and in the revisions of this manuscript.

The contents of this publication are the sole responsibility of the authors. The content of this publication does not necessarily reflect the views, opinions, or policies of the NIH, the Uniformed Services University of the Health Sciences, the US Department of Health and Human Services, the US Department of Defense, or the US Government, nor does mention of trade names, commercial products, or organizations imply endorsement by the US government. This work was prepared by a military or civilian employee of the US Government as part of the individual’s official duties. Therefore, it is in the public domain and does not possess copyright protection. Public domain information may be freely distributed and copied; however, as a courtesy, it is requested that the authors be given an appropriate acknowledgement.

This work was supported by the National Human Genome Research Program Intramural Research Program, grant numbers: ZIA HG200417, ZIC HG200420; and the Division of Intramural Research of the National Institute of Allergy and Infectious Diseases. The All of Us Research Program is supported by the National Institutes of Health, Office of the Director: Regional Medical Centers: 1 OT2 OD026549; 1 OT2 OD026554; 1 OT2 OD026557; 1 OT2 OD026556; 1 OT2 OD026550; 1 OT2 OD 026552; 1 OT2 OD026553; 1 OT2 OD026548; 1 OT2 OD026551; 1 OT2 OD026555; IAA #: AOD 16037; Federally Qualified Health Centers: HHSN 263201600085U; Data and Research Center: 5 U2C OD023196; Biobank: 1 U24 OD023121; The Participant Center: U24 OD023176; Participant Technology Systems Center: 1 U24 OD023163; Communications and Engagement: 3 OT2 OD023205; 3 OT2 OD023206; and Community Partners: 1 OT2 OD025277; 3 OT2 OD025315; 1 OT2 OD025337; 1 OT2 OD025276. Funders played no role in study design, data collection, analysis and interpretation of data, or the writing of this manuscript.

## AUTHOR CONTRIBUTIONS

BJW and JCD conceived of the study design. BJW constructed the phenotype, conducted all analyses, and drafted the manuscript. TCT and HM contributed foundational data analysis support. FABC and EER offered appraisal of data analysis and early revision of the manuscript. All authors contributed to the final revision of the manuscript.

