## Supplementary Figures for "Computable Phenotypes for Respiratory Viral Infections in the *All of Us* Research Program"

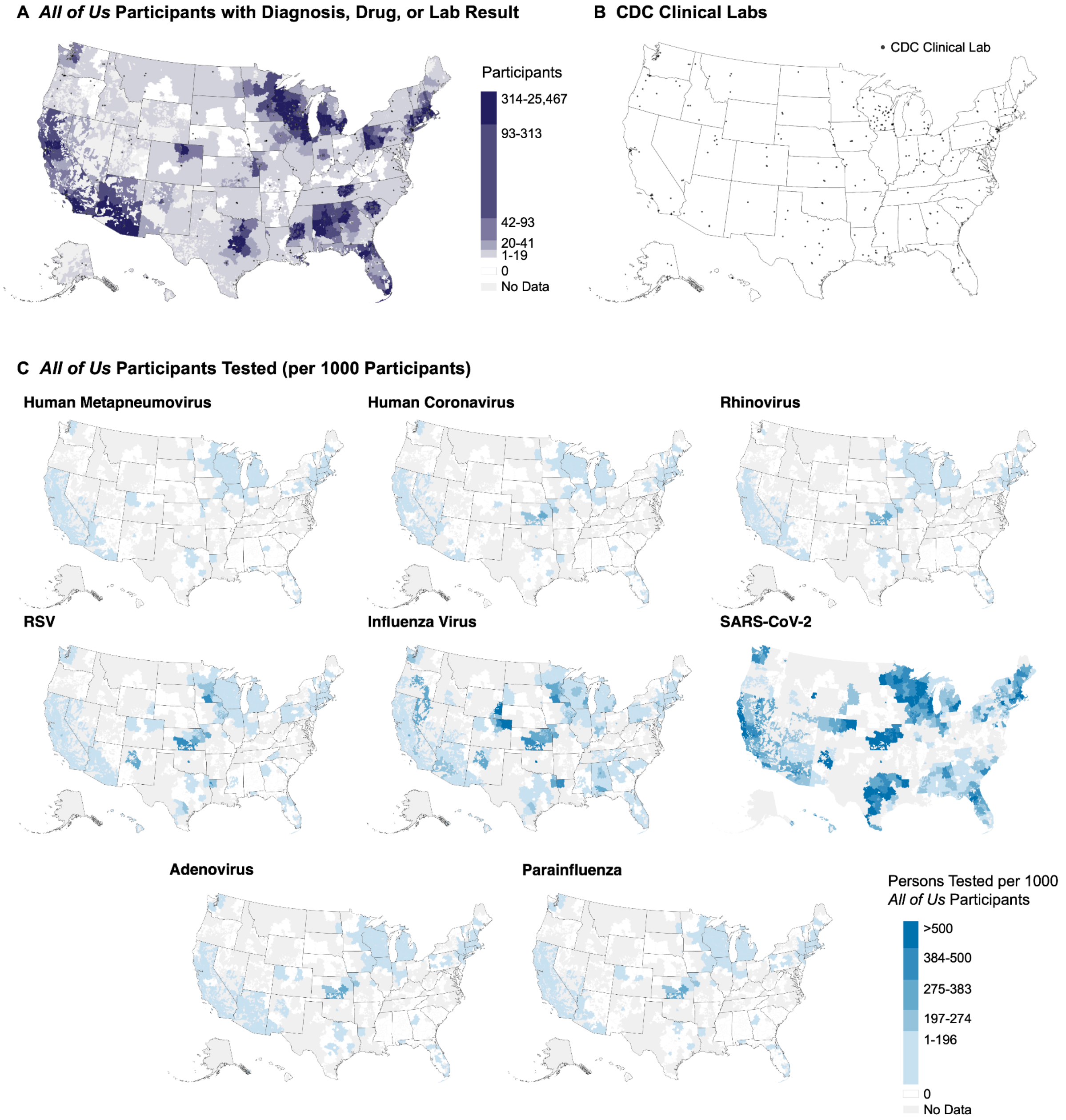


**Supplementary Figure 1.** Geographic comparison of testing data. (A) Total *All of Us* participants with a diagnosis, drug prescription, or laboratory test result per three-digit zip code prefix, color-coded by quintile. (B) Locations of CDC clinical labs reporting to NREVSS. (C) *All of Us* testing rates per 1,000 *All of Us* participants with EHR data by zip3, colored using SARS-CoV-2-defined quintiles to enable comparisons across testing rates of pathogens. Three-digit zip code regions ≤ 5 *All of Us* participants with EHR data were marked as “No Data” in (C). RSV: respiratory syncytial virus.

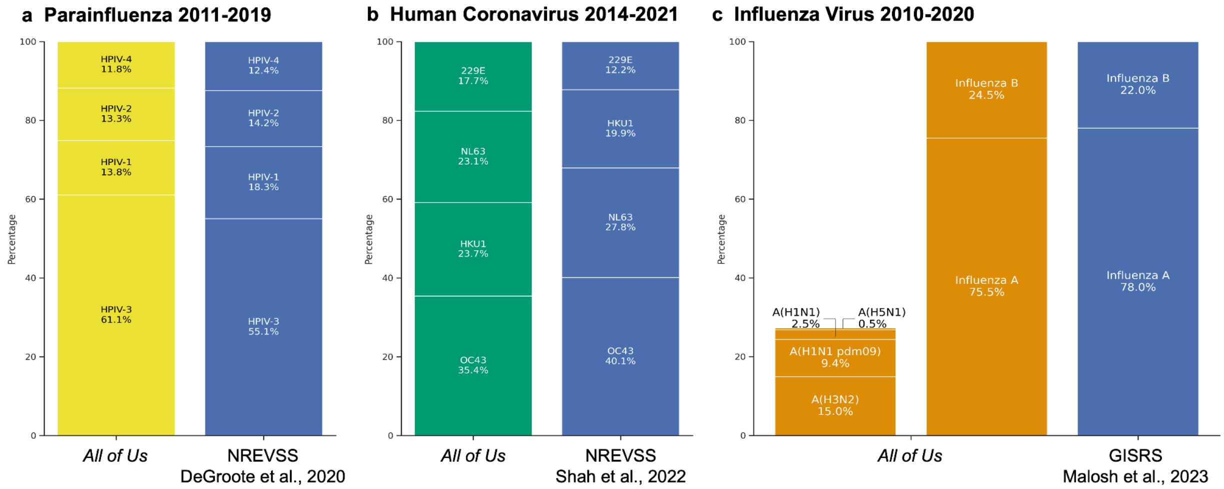


**Supplementary Figure 2.** Comparison of *All of Us* virus type or subtype distributions with national surveillance data. Comparison of (A) parainfluenza (HPIV) types from NREVSS (July 1, 2011 to June 30, 2019), (B) human coronavirus (hCoV) types from NREVSS (July 5, 2014 to November 6, 2021), and (C) influenza virus types (also shown with subtypes) from GISRS (2010-2020) against *All of Us* data from corresponding time periods. While GISRS influenza data were limited to epidemiologic seasons, *All of Us* data incorporated year-round results.

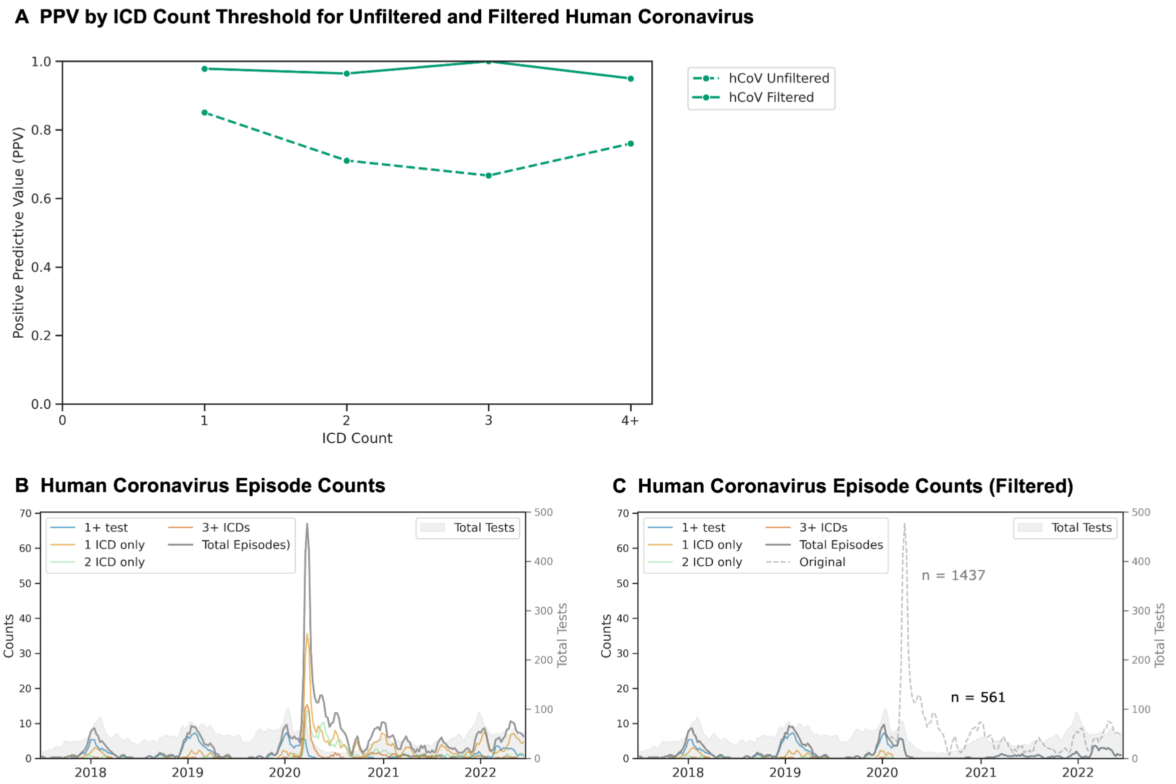


**Supplementary Figure 3.** Impact of COVID-19 pandemic on human coronavirus episodes. (A) PPV of ICD codes using laboratory results as the reference standard for human coronavirus (hCoV), before (dashed line) and after (solid line) removing hCoV ICD codes after February 1, 2020. Three-week moving averages for hCoV showing total episodes (gray), episode components (colored lines), and testing volume (gray shading) before (B) and after (C) filtering. For B and C, component types include positive tests (blue), ICD codes only (light orange: 1, light green: 2, red-orange: ≥ 3), and medications (dark green: online, pink: with ICD codes). The left y-axis corresponds to episodes, and the right y-axis corresponds to test volume.

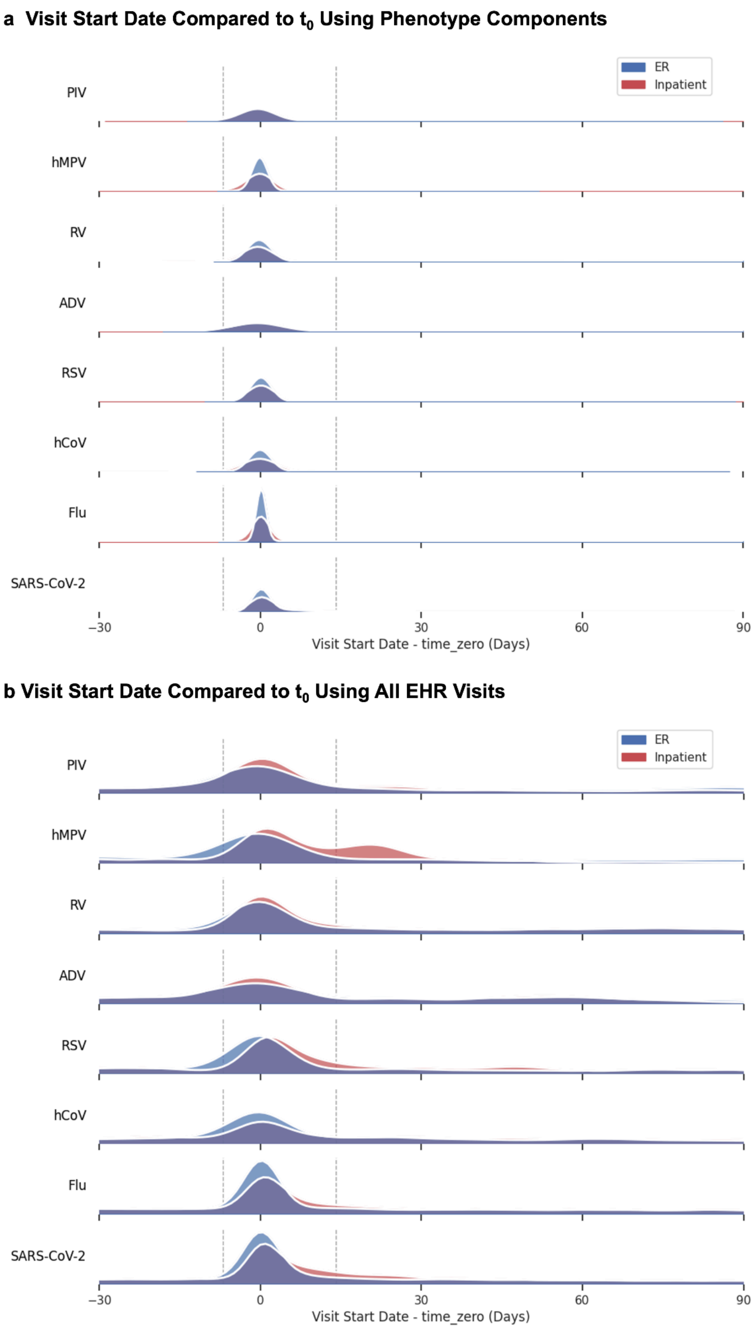


**Supplementary Figure 4**: Temporal distribution of visits relative to episode $t_0$. Kernel density estimates showing visit timing relative to $t_0$ for (A) visits associated with phenotyped components and (B) all ER and inpatient visits from -60 to +120 days. dashed lines indicate the -7 to +14 day window used for level of care acuity assessment. ER: emergency room; PIV: parainfluenza; hMPV: human metapneumovirus; RV: rhinovirus; ADV: adenovirus; RSV: respiratory syncytial virus; hCoV: human coronavirus; Flu: influenza virus.

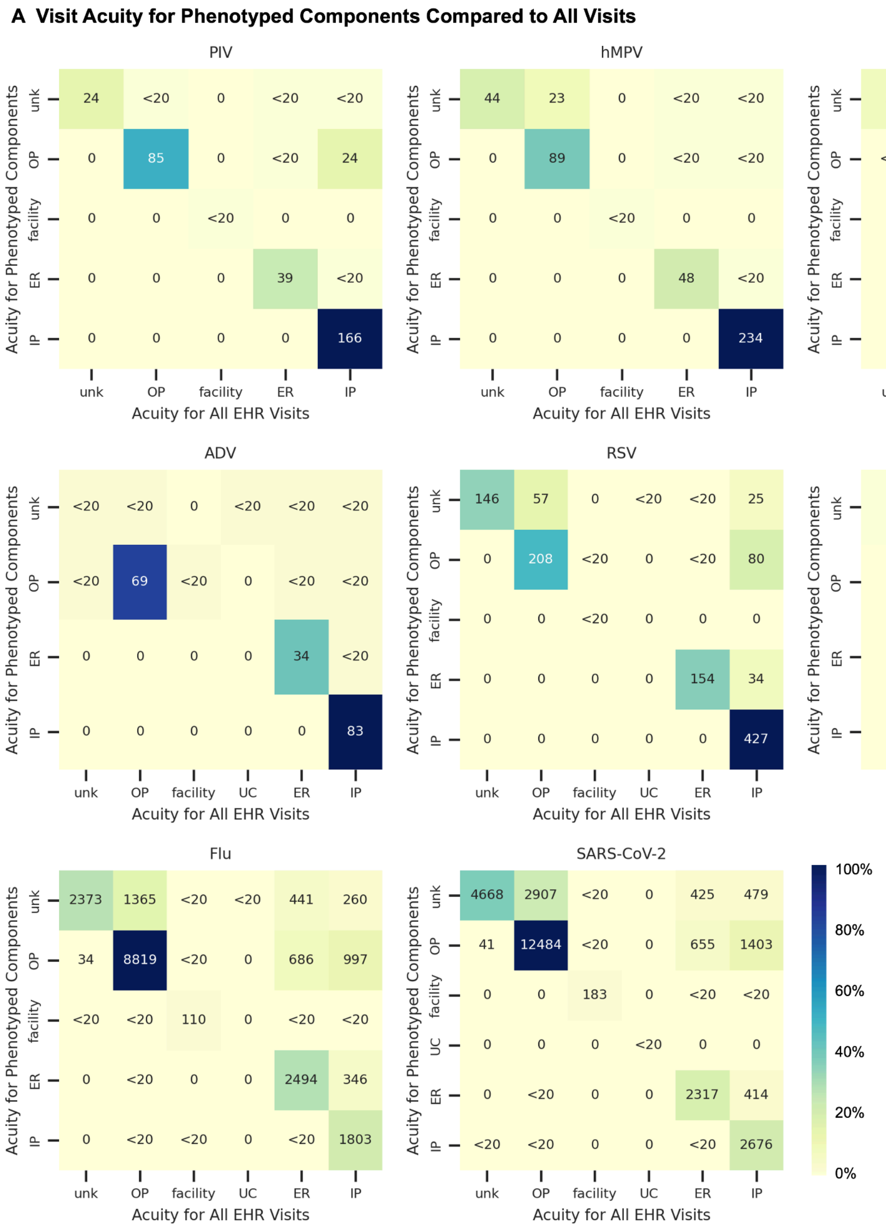


**Supplementary Figure 5**: Comparison of level of care sources. Maximum level of care acuity from phenotype-related visits (rows) versus all visits in the -7 to +14 day window (columns). IP: inpatient; ER: emergency room; UC: urgent care; OP: outpatient; Unk: unknown; PIV: parainfluenza; hMPV: human metapneumovirus; RV: rhinovirus; ADV: adenovirus; RSV: respiratory syncytial virus; hCoV: human coronavirus; Flu: influenza virus.

### CODE-EHR framework: Best practice checklist to report on the use of structured electronic healthcare records in clinical research

Date of completion: Study name: All of Us Respiratory Virus Computable Phenotype

| Item | Objective | Framework standards | Minimum information to provide | Lead Author acknowledgement |
|---|---|---|---|---|
| 1. Dataset construction and linkage | To provide an understanding of how the structured healthcare data were identified and used. | Minimum: Flow diagram of datasets used in the study, and description of the processes and directionality of any linkage performed, published within the research report or supplementary documents.<br>Preferred: Provided within a pre-published protocol or open-access document. | (a) State the source of any datasets used.<br>(b) Comment on how the observed and any missing data were identified and addressed, and the proportion observed for each variable.<br>(c) Provide data on completeness of follow-up.<br>(d) For linked datasets, specify how linkage was performed and the quality of linkage methods. | Select one option:<br>Minimum standard not met ☐<br>Minimum standard met ☐<br>Preferred standard met ☒ |
| 2. Data fit for purpose | To ensure transparency with the approach taken, with respect to coding of the structured healthcare data. | Minimum: Clear unambiguous statements on the process of coding in the methods section of the research report.<br>Preferred: Provided within a pre-published protocol or open-access document. | (a) Confirm origin, clinical processes, and the purpose of data.<br>(b) Specify coding systems, clinical terminologies or classification used and their versions, and any manipulation of the coded data.<br>(c) Provide detail on quality assessment for data capture.<br>(d) Outline potential sources of bias. | Select one option:<br>Minimum standard not met ☐<br>Minimum standard met ☒<br>Preferred standard met ☐ |
| 3. Disease and outcome definitions | To fully detail how conditions AND outcome events were defined, allowing other researchers to identify errors and repeat the process in other datasets. | Minimum: State what codes were used to define diseases, treatments, conditions and outcomes *prior to statistical analysis*, including those relating to patient identification, therapy, procedures, comorbidities, and components of any composite endpoints.<br>Preferred: Provided within a pre-published protocol or open-access document *prior to statistical analysis.* | (a) Detailed lists of codes used for each aspect of the study.<br>(b) Date of publication and access details for the coding manual (please add to box below).<br>(c) Provide definitions, implementation logic and validation of any phenotyping algorithms used.<br>(d) Specify any processes used to validate the coding scheme or reference to prior work. | Select one option:<br>Minimum standard not met ☐<br>Minimum standard met ☒<br>Preferred standard met ☐ |
| 4. Analysis | To fully detail how outcome events were analysed and allow independent assessment of the authenticity of study findings. | Minimum: Describe the process used to analyse study outcomes, including statistical methods and use of any machine learning or algorithmic approaches.<br>Preferred: Provide a statistical analysis plan as a supplementary file, locked prior to analyses commencing. | (a) Provide details on all statistical methods used.<br>(b) Provide links to any machine code or algorithms used in the analysis, preferably as open-source.<br>(c) Specify the processes of testing assumptions, assessing model fit and any internal validation.<br>(d) Specify how generalisability of results was assessed, the replication of findings in other datasets, or any external validation. | Select one option:<br>Minimum standard not met ☐<br>Minimum standard met ☒<br>Preferred standard met ☐ |

| | | | | |
|---|---|---|---|---|
| 5. Ethics and governance | To provide patients, who may or may not have given consent, and regulatory authorities the ability to interrogate the security and provenance of the data. | Minimum: Clear unambiguous statements on how the principles of Good Clinical Practice and Data Protection will be/were met, provided in the methods section of the research report.<br>Preferred: Provided within a pre-published protocol or open-access document with evidence of patient and public engagement. | (a) State how informed consent was acquired, or governance if no patient consent.<br>(b) Specify how data privacy was protected in the collection and storage of data.<br>(c) Detail what steps were taken for patient and public involvement in the research study.<br>(d) Provide information on where anonymised source data or code can be obtained for verification and further research. | Select one option:<br>Minimum standard not met ☐<br>Minimum standard met ☒<br>Preferred standard met ☐ |
| 6. Coding manual | DOI of publication or website address: workbench.researchallofus.org/workspaces/aou-rw-ae307fda/respiratoryviralinfectionsinallofus/analysis<br>Date published: 1/15/25 | | | |
| 7. Comments | | | | |
| **8. Summary declaration** | One or more minimum standards not met ☐ OR **All minimum standards met** ☒<br>Number of preferred standards met: 1 / 5 | | | |

Directions for use:

**Research team:** To complete the checklist, authors will need to consider these points during the design of the research to ensure that coding protocols and coding manuals are pre-published. Where applicable, it is advisable that all five minimum standards are met for an individual research study, whether observational or a controlled trial. If any component is not applicable to the study, the corresponding author can indicate why this is the case in the comment box. This checklist can accompany the article as a supplementary file on submission to the journal, with the ability for readers to review responses. A comment on the meeting of standards in the text of the method section is suggested, for example; *"this study meets all five of the CODE-EHR minimum framework standards for the use of structured healthcare data in clinical research, with two out of five standards meeting preferred criteria <add reference to this CODE-EHR paper; https://doi.org/10.1136/bmj-2021-069048>"; OR "this study meets four out of five of the CODE-EHR minimum framework standards for the use of structured healthcare data in clinical research; one of the five minimum standards was not met as coding schemes were not specified prior to analysis <add reference to this CODE-EHR paper; https://doi.org/10.1136/bmj-2021-069048>." Note, easy to complete form versions of this checklist are available in the article appendices (word and pdf versions) and at https://www.escardio.org/bigdata.*

**Research appraisers (patients, clinicians, regulators, guideline task forces):** Where applicable, it is advisable that all five minimum standards are met for the research study to be considered robust.

**FURTHER DETAILS ON THE CODE-EHR FRAMEWORK:** please refer to Kotecha D, Asselbergs FW, et al; on behalf of the Innovative Medicines Initiative BigData@Heart Consortium, European Society of Cardiology, CODE-EHR international consensus group. CODE-EHR best practice framework for the use of structured electronic healthcare records in clinical research. BMJ 2022;378:e069048. doi:10.1136/bmj-2021-069048. Also published in Lancet Digit Health and Eur Heart J.
